# Stratification of Pediatric COVID-19 cases by inflammatory biomarker profiling and machine learning

**DOI:** 10.1101/2023.04.04.23288117

**Authors:** Devika Subramanian, Aadith Vittala, Xinpu Chen, Christopher Julien, Sebastian Acosta, Craig Rusin, Carl Allen, Nicholas Rider, Zbigniew Starosolski, Ananth Annapragada, Sridevi Devaraj

## Abstract

An objective method to identify imminent or current Multi-Inflammatory Syndrome in Children (MIS-C) infected with SARS-CoV-2 is highly desirable. The aims was to define an algorithmically interpreted novel cytokine/chemokine assay panel providing such an objective classification. This study was conducted on 4 groups of patients seen at multiple sites of Texas Children’s Hospital, Houston, TX who consented to provide blood samples to our COVID-19 Biorepository. Standard laboratory markers of inflammation and a novel cytokine/chemokine array were measured in blood samples of all patients. Group 1 consisted of 72 COVID-19, 66 MIS-C and 63 uninfected control patients seen between May 2020 and January 2021 and predominantly infected with pre-alpha variants. Group 2 consisted of 29 COVID-19 and 43 MIS-C patients seen between January-May 2021 infected predominantly with the alpha variant. Group 3 consisted of 30 COVID-19 and 32 MIS-C patients seen between August-October 2021 infected with alpha and/or delta variants. Group 4 consisted of 20 COVID-19 and 46 MIS-C patients seen between October 2021-January 2022 infected with delta and/or omicron variants. Group 1 was used to train a L1-regularized logistic regression model which was validated using 5-fold cross validation, and then separately validated against the remaining naïve groups. The area under receiver operating curve (AUROC) and F1-score were used to quantify the performance of the algorithmically interpreted cytokine/chemokine assay panel. Standard laboratory markers predict MIS-C with a 5-fold cross-validated AUROC of 0.86 ± 0.05 and an F1 score of 0.78 ± 0.07, while the cytokine/chemokine panel predicted MIS-C with a 5-fold cross-validated AUROC of 0.95 ± 0.02 and an F1 score of 0.91 ± 0.04, with only sixteen of the forty-five cytokines/chemokines sufficient to achieve this performance. Tested on Group 2 the cytokine/chemokine panel yielded AUROC =0.98, F1=0.93, on Group 3 it yielded AUROC=0.89, F1 = 0.89, and on Group 4 AUROC= 0.99, F1= 0.97). Adding standard laboratory markers to the cytokine/chemokine panel did not improve performance. A top-10 subset of these 16 cytokines achieves equivalent performance on the validation data sets. Our findings demonstrate that a sixteen-cytokine/chemokine panel as well as the top ten subset provides a sensitive, specific method to identify MIS-C in patients infected with SARS-CoV-2 of all the major variants identified to date.

## Introduction

Severe Acute Respiratory Syndrome Coronavirus 2 (SARS-CoV-2) infection, is typically milder in children than in adults; yet a significant number of patients still need hospitalization (1). A life-threatening consequence of the infection in children is multisystem inflammatory syndrome (MIS-C). The WHO definition of MIS-C describes this condition occurring in patients under 19 years of age, presenting within 12 weeks following SARS CoV-2 primary infection or exposure with high fever for at least 3 days, two or more clinical features, disease activity and at least one elevated laboratory marker of inflammation (2). While the risk factors that predispose some children to develop MIS-C are not fully understood, identification of MIS-C is important because severe organ dysfunction and death have been reported in these patients (3, 4). A prediction of MIS-C at initial presentation enables early initiation of immunotherapies that reduce severity and improve outcomes. Recent studies in adults (5) suggest that the cytokine storm in SARS CoV-2 infected patients is directly linked to disease severity. In this study, we therefore aimed to characterize the cytokine/chemokine profile of pediatric patients with MIS-C compared to those with SARS-CoV-2 infection (COVID-19) and to derive a method to stratify pediatric patients presenting with COVID-19 by their risk of MIS-C.

## Subjects and Methods

### Study design

Serum and plasma samples were obtained from the Texas Children’s Hospital COVID-19 Biorepository (TCB) established in April 2020 under a protocol approved by the Institutional Review Board at Baylor College of Medicine. This repository holds serum/plasma samples from patients admitted with COVID-19 and/or MIS-C to any of the sites of the Texas Children’s Hospital (TCH) system. In addition, samples from patients with no known inflammatory condition were included as controls. The TCH system serves the greater Houston metropolitan area of about 10,000 square miles, which incorporates 9 counties with a combined population of 8.2 million. This study used a training cohort of 201 patients who presented at one of the sites of TCH between April 2020 and January 2021. Of this cohort, 66 had a diagnosis of MIS-C by the CDC criteria, 72 were diagnosed with COVID-19 but did not develop MIS-C, and 63 were controls. The training cohort was obtained when the pre-alpha strain of the SARS-Cov-2 virus was predominant in the population. Three additional validation sets were also obtained. Validation cohort 1 constituted 92 new patients (43 MIS-C, 29 COVID-19, 20 controls) treated in the TCH system between January 2021 and May 2021. Validation cohort 2 constituted 78 new patients (32 MIS-C, 30 COVID-19, 16 controls) treated in the TCH system between August 2021 and October 2021. Validation cohort 3 had 76 patients (20 COVID-19, 46 MIS-C, 10 controls) treated in the TCH system between October 2021 and January 2022. While the variants infecting the specific patients from whom these samples were drawn were not recorded, the distribution of strains in TCH system patients as a function of time was recorded (Supplementary Figure 1a and 1b).

Based on this temporal distribution we infer that the training cohort were infected with predominantly pre-alpha variants, validation cohort 1 predominantly alpha, validation cohort 2 predominantly delta and validation cohort 3 predominantly delta and omicron variants of SARS-CoV-2. Supplementary Figure 2 shows the distribution of time intervals between a patient receiving diagnosis of MIS-C and the time of blood sampling, where negative times indicate that the sample was drawn after the diagnosis had already been made, while positive times indicate that the patient received a diagnosis of MIS-C after the blood sample was drawn. Of note roughly half of the MIS-C patients had not yet met CDC criteria nor had they received a diagnosis at the time the sample was drawn.

## Methods

### Serum protein/laboratory data/EHR data collection

Frozen aliquots of serum/plasma from the TCB were used for biomarker analysis. Values of thirteen clinically used laboratory markers of disease activity and inflammation (C-Reactive Protein (CRP), Procalcitonin, D-Dimer, B-type Natriuretic Peptide (BNP), Sodium, Platelet count, Albumin, Fibrinogen, Protime, Neutrophil to Lymphocyte ratio (NLR), Total CO_2_, Ferritin and Troponin I) were measured since they have been shown previously to be associated with both adult and pediatric SARS-CoV-2 infection (5, 6). These laboratory markers were measured on blood samples collected at approximately the same time as the biorepository samples. Demographic data (age, gender, race, ethnicity), vitals, and results of SARS-CoV-2 antibody and PCR testing were all gathered from the patients’ electronic health records (EHR). In addition, specifics of treatments administered (IVIG, anti-IL1RA and steroids, etc.) during the hospital stay were gathered from the EHR. Parameters marking the severity of disease, including length of stay in the hospital, length of stay in the ICU, and use of ventilators, ECMO, oxygen and CPAP were all also obtained from the EHR.

### Confirmation of SARS-CoV-2 infection

All patients whose nasopharyngeal swab tested positive for SARS-CoV-2 using either a transcription-mediated amplification assay or reverse-transcriptase PCR assay were considered confirmed cases of SARS-CoV-2 infection. For MIS-C clinical definition, the CDC criteria were applied.

### Cytokine/Chemokine Profiling

Cytokines/chemokines were analyzed using a 48-plex Millipore MAP Human Cytokine/Chemokine Magnetic Bead Panel (Millipore). Each sample was run in duplicate in separate measurements on a Luminex® MAGPIX instrument. Both kit-derived quality controls (QCs) and an in-house sample pool were used to control for lot-to-lot variability. 45 of the 48 cytokines/chemokines with less than 10% inter- and intra-assay coefficients of variation, and with less than 15% difference in duplicate readings were retained for analysis. The full list of cytokines/chemokines selected for the training cohort as well as the three validation cohorts is shown in Table 1c.

### Computation

All calculations and computation were performed in, and all algorithms implemented in, Anaconda Python 3.9 with the sklearn and scipy packages.

### Univariate discrimination by the Wilcoxon-rank-sum test

The ability of laboratory and cytokine/chemokine analytes to discriminate COVID-19 from MIS-C groups was assessed using the non-parametric Wilcoxon-rank-sum test on the training data (72 COVID-19 samples, 66 MIS-C samples). We tested the null hypothesis that there is no difference in the medians of the biomarker values between the COVID-19 and MIS-C cohorts. p ≤ 0.05 was used to indicate significant differences in the distributions of the analyte in the two populations.

### Multivariate cross-validated L1 regularized logistic regression classification

An L1-regularized logistic regression augmenting the cross-entropy loss function with a penalty term proportional to the sum of the absolute values of the regression coefficients (7, 8) was used for supervised learning. 5-fold cross validation was used to ensure generalization performance within the training set, characterized by the Area Under the Receiver Operating Curve (AUROC) and the F1-score (9).

### Multidimensional data representation

The Uniform MAnifold expression and Projection (10-12) (UMAP) technique was used to reduce dimensionality of the dataset and visualize multidimensional cytokine/chemokine data in an unsupervised manner.

### Network analysis

The STRING database (13) of protein-protein interactions including both physical associations and functional associations was used to identify protein interaction networks associated with the cytokine/chemokine combinations characteristic of COVID-19 and MIS-C. Interactions with confidence score > 0.9 were selected for network construction. In addition, we added connections between any two cytokines/chemokines that had high correlations (Pearson correlation > 0.8) in our training data. To visualize networks of proteins affected by either COVID-19 or MIS-C, we computed subnetworks composed of all cytokines elevated or depressed two-fold or more when comparing the median level for patients with either COVID-19 or MIS-C to the levels in control patients. To place the subnetworks in a more global context, we also added proteins that were not measured into the network if they interacted with at least three measured cytokines/chemokines in the network. Graph construction and visualization were done using custom-written Python 3.9 scripts.

## Results

### Clinical characteristics of the training cohort and validation sets

Table 1a summarizes clinical and demographic characteristics for the training cohort, and results of the Wilcoxon rank-sum test testing the null hypothesis that there is no difference in each variable between MIS-C and COVID-19 patients. Table 1b summarizes the same characteristics for validation sets 1, 2 and 3. In the training cohort, significant differences between the MIS-C and COVID-19 groups were noted in median age (9 versus 14 years), sex (62% male versus 46%), median hospital stay duration (7.8 days versus 5.5 days), median ICU stay (3.4 days versus 0 days), ventilator use (30.8% vs 17.1%), and incidence of acute kidney injury (AKI) (35.3% versus 11.4%). There were no significant differences in the distribution of race, ethnicity, and BMI values in the two cohorts. The demographics of the three validation cohorts mirror that of the training set; except for the COVID- 19 cohort of validation set 3 which has only 20 patients. The differences between the COVID-19 and MIS-C cohorts in median hospital stay duration, median ICU stay, ventilator use, and AKI that are observed in the training cohort, are preserved in the three validation sets. However, the median values of these quantities decrease over time which is consistent with evolution of the virus to higher infectivity/lower disease severity, as well as improvements in disease management and use of vaccines prior to validation set 3. For example, the median length of stay in hospital for the COVID-19 group goes down from 5.5 days in the training cohort to 3.8 days in validation sets 1 and 2 to 0.5 days in validation set 3. A corresponding drop is observed in the length of hospital stay for the MIS-C cohort starting at 7.8 days through to 6.7 days, 6.0 days, to 5.8 days in validation set 3. Similar drops over time are seen for median ICU length of stay, ventilator use, CPAP, and AKI in both MIS-C and COVID-19 cohorts in the three validation sets.

**Table 1.** **a:** Clinical, demographic and laboratory characteristics of training cohort of 72 COVID and 66 MIS-C patients and validation cohort 1 (29 COVID-19, 43 MIS-C) and cohort 2 (30 COVID-19, 32 MIS-C). Categorical variables are expressed as frequencies or percentages, while continuous quantities are represented as median/interquartile range pairs. The variables that show significant differences between the COVID-19 and MIS-C cohorts at p < 0.05, using the Wilcoxon-rank-sum test for continuous quantities and the chi-squared test for discrete quantities, are in italics.

|  | **COVID-19**<br>**Training (N=72)** | **MIS-C**<br>**Training (N=66)** | **p-value** |
| --- | --- | --- | --- |
| **Age** at sample collection Med/IQR (yrs) | 14 (6-18) | 9 (6-14) | *0.013* |
| **BMI** Med/IQR | 20.8 (17.6-30.5) | 19.4 (17.1-25.8) | 0.4 |
| **Gender** |  |  | *0.03* |
| M/F % | 46/54 | 62/38 |  |
| **Race** |  |  | 0.95 |
| White % | 65.7 | 63.7 |  |
| African American % | 28.7 | 28.9 |  |
| Asian % | 2.8 | 4.3 |  |
| Other % | 2.8 | 2.9 |  |
| **Ethnicity** |  |  | 0.65 |
| Hispanic % | 54 | 49 |  |
| not Hispanic % | 46 | 51 |  |
| **SARS-CoV-2 test** |  |  | *$1.3 \times 10^{-11}$* |
| PCR % | 90 | 0 |  |
| Antibody % | 10 | 100 |  |
| unknown % | 0 | 0 |  |
| **IVlg/steroids** 7dys before collection. % | 33 | 46 | 0.08 |
| **IVlg/steroid from collection** days | -8.5 (-41, -1) | 0 (-2.75,1) | *$4.9 \times 10^{-10}$* |
| **LOS in hospital** day | 5.5 (2.8,12.3) | 7.8 (5.3,11.5) | *0.03* |
| **ICU LOS** days | 0 (0,4.5) | 3.4 (1.5,6.7) | *0.002* |
| **ECMO** % | 1.4 | 4.6 | 0.36 |
| **Ventilator** % | 17.1 | 30.8 | *0.035* |
| **CPAP** % | 15.7 | 24.6 | 0.16 |
| **AKI** % | 11.4 | 35.3 | *$1.3 \times 10^{-3}$* |
| **Labs** |  |  |  |
| Sodium mEq/L | 138 (136,140) | 134 (131,137) | *$2.5 \times 10^{-7}$* |
| Albumin g/dL | 4 (3.4,4.6) | 3.3 (3.0,3.7) | *$9.7 \times 10^{-7}$* |
| CO2 mEq/L | 26(23,28) | 24(20,26) | *$3.8 \times 10^{-3}$* |
| BNP pg/mL | 64.1(18.5,67.5) | 221.6(49.8,589.2) | *$1.5 \times 10^{-7}$* |
| Tropl ng/mL | 0.02(0.01,0.07) | 0.02(0.01,0.08) | 0.12 |
| Platelets #/nL | 280(176,404) | 158(123,240) | *$7.0 \times 10^{-7}$* |
| Protime seconds | 14.6(14,14.9) | 15.4(14.6,16.5) | *$1.6 \times 10^{-4}$* |
| D-dimer μg/mL | 1.1(0.5,2.04) | 3.0(1.7,4.06) | *$1.4 \times 10^{-7}$* |
| Fibrinogen mg/dL | 402(317,459) | 513(424,600) | *$1.8 \times 10^{-5}$* |
| Procalcitonin ng/mL | 1.6 (0.1,2.1) | 4.7 (1.6,12.8) | *$4.6 \times 10^{-9}$* |
| CRP mg/dL | 2.1 (0.5,5.7) | 15.9(5.2,24.3) | *$5.2 \times 10^{-10}$* |
| NLRatio | 2.5(1.3,5.4) | 8(3.1,14.6) | *$3.3 \times 10^{-4}$* |
| Ferritin ng/mL | 254(97,377) | 333(166,641) | *$7.2 \times 10^{-3}$* |

**Table 1.**
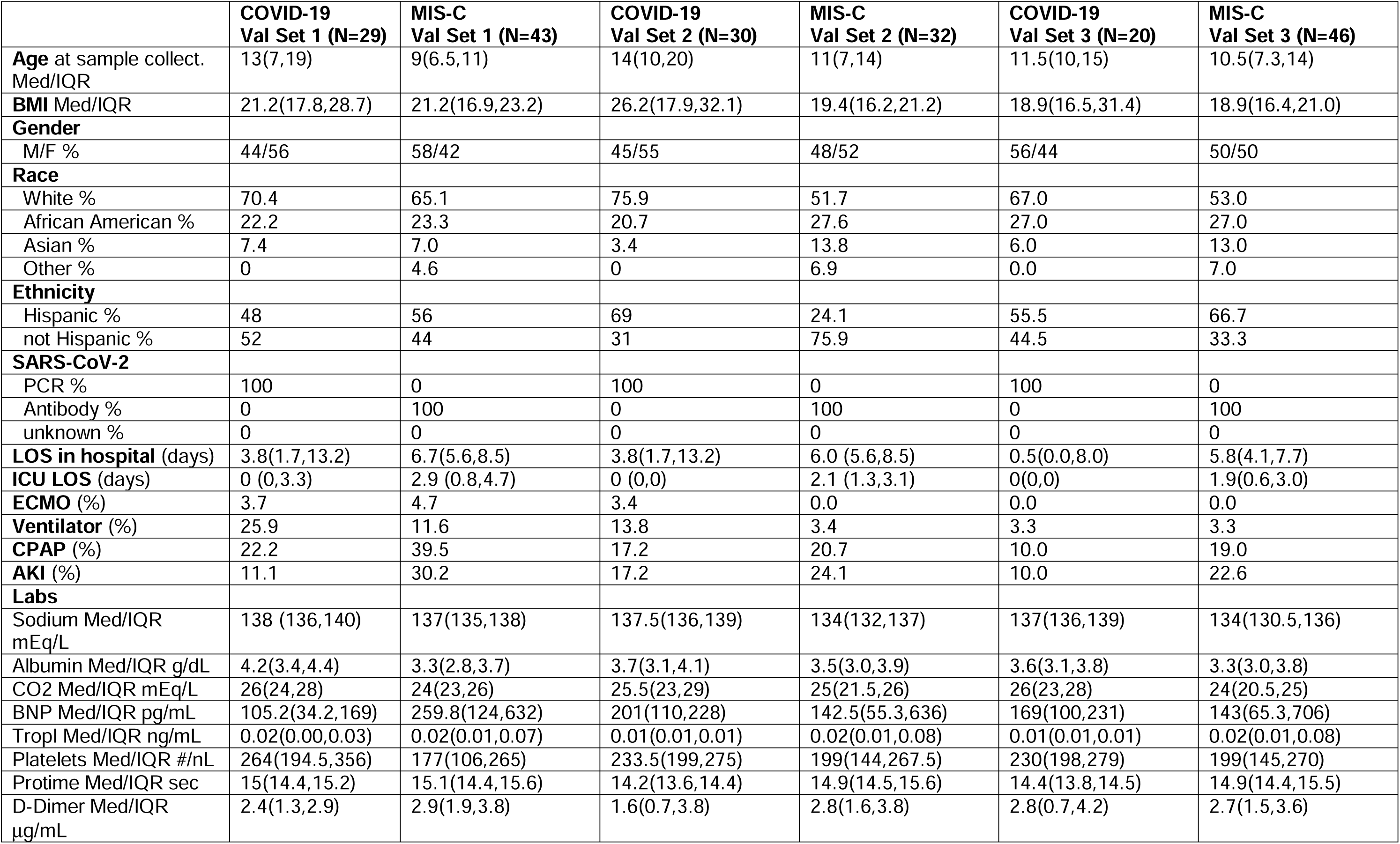
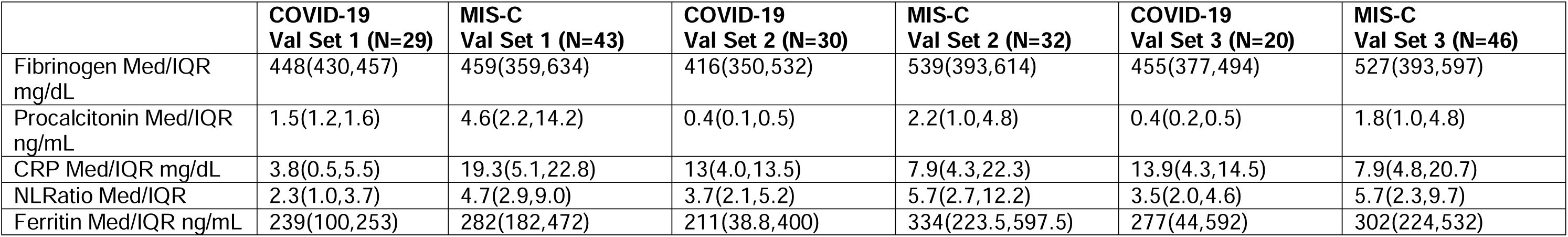
**b:** Clinical, demographic and laboratory characteristics of the validation cohort 1 (29 COVID-19, 43 MIS-C), validation cohort 2 (30 COVID-19, 32 MIS-C) and validation cohort 3 (20 COVID-19, 46 MIS-C). Categorical variables are expressed as frequencies or percentages, while continuous quantities are represented as median/interquartile range pairs.

### Laboratory characteristics of the training cohort and validation sets

Table 1a also summarizes statistics for all the measured laboratory markers for the training cohort, and Table 1b provides these statistics for validation sets 1, 2 and 3. Markers that were significantly elevated in the MIS-C training cohort compared to the COVID-19 training cohort are: CRP (median 15.9 mg/dL, p=5.2 X 10^-10^), ferritin (median 333.3 mg/L, p=7.3 X 10^-3^) and procalcitonin (median 4.7 ng/mL, p=4.6 X 10^-9^), fibrinogen (median 503 mg/dL, p=1.8 X 10^-5^), protime (median 15.4 s, p=1.6 X 10^-4^), D-dimer (median 93.0 µg/mL, p=1.4 X 10^-7^), BNP (median 221.6 pg/mL, p=1.5 X 10^-7^) and the neutrophil to lymphocyte ratio (NLR), (median 8.0, p=3.4 X 10^-4^). Markers that were significantly lowered in the MIS-C training cohort compared to the COVID-19 training cohort are albumin (median 3.3 g/dL, p=9.7 X 10^-7^), platelet counts (median 158/µL, p=7.0 X 10^-7^) sodium (median 134, p=2.1 X 10^-7^). Troponin I level between the two cohorts were not significantly different.

In the three validation sets, for the MIS-C cohort, ferritin, fibrinogen, protime, D-Dimer, albumin and sodium levels remain about the same as in the training set. However, other inflammation markers improve from the training MIS-C cohort (pre-alpha) to the MIS-C cohort in validation set 3 (delta and omicron variants): platelet counts increase from 158/µL in the training MIS-C cohort to 177/µL in validation set 3, CRP declines from 15.9 mg/dL to 7.9 mg/dL, procalcitonin drops from 4.7 ng/mL to 1.8 ng/mL, and BNP decreases from 221.6 pg/mL to 143 pg/mL. In the three validation sets, for the COVID-19 cohort, there are significant changes in four laboratory markers of inflammation from the training group to validation set 3: albumin drops from 4.2 g/dL to 3.6 g/dL (closer to the MIS-C group in the training set), BNP increases from 64 pg/mL to 169 pg/mL, procalcitonin from 1.6 to 0.4, and CRP from 2.1 mg/dL to 13.9 mg/dL. These data are consistent with MIS-C cases getting milder as the virus evolves, and COVID-19 becoming more severe, relative to the initial pre-alpha manifestation. Further, these trends suggest that standard laboratory markers of inflammation change with the severity of the variant/disease course an d cannot distinguish between MIS-C and COVID-19 reliably.

### Cytokine/chemokine profiles of the training cohort

Table 1c shows the levels of 45 cytokines/chemokines ordered by the value of p from the Wilcoxon-rank-sum univariate test differentiating COVID-19 from MIS-C in the training cohort. 34 of the 45 cytokines/chemokines were statistically significantly different in the univariate sense. The top sixteen markers of these are soluble IL2 receptor, IP-10, MIG, IL-10, IL-15, IL-3, IL-1RA, TNF-α, IL-13, IFN-1, IL-22, IL-2, TGF, GCSF, IL-6, and IL-27 all with p <3x10^-6^. Also shown in Table 1c are the specificity and sensitivity of each individual biomarker with 5-fold cross-validation on the training data. No single cytokine/chemokine has both specificity and sensitivity over 0.9.

**Table 1.**
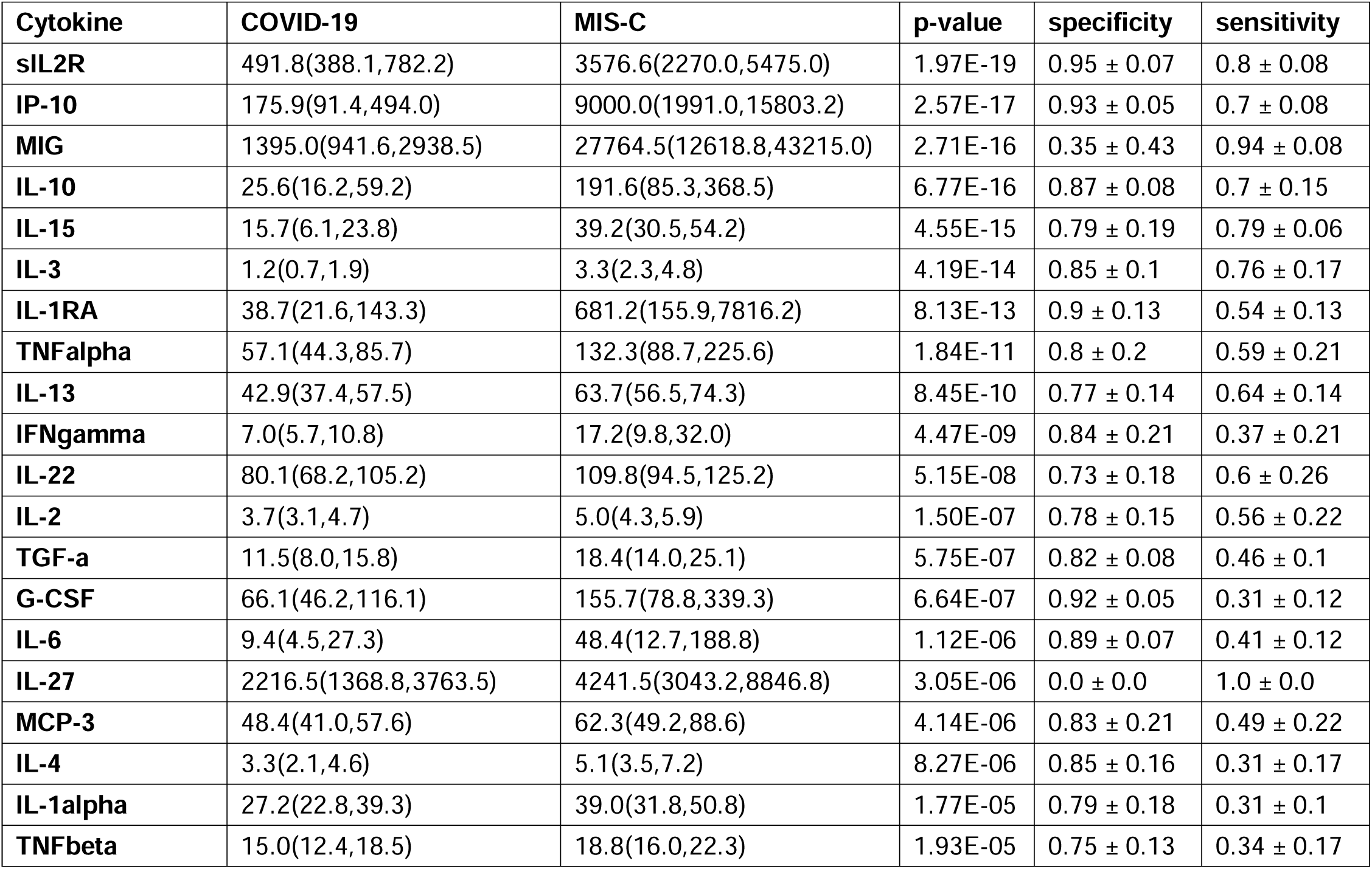
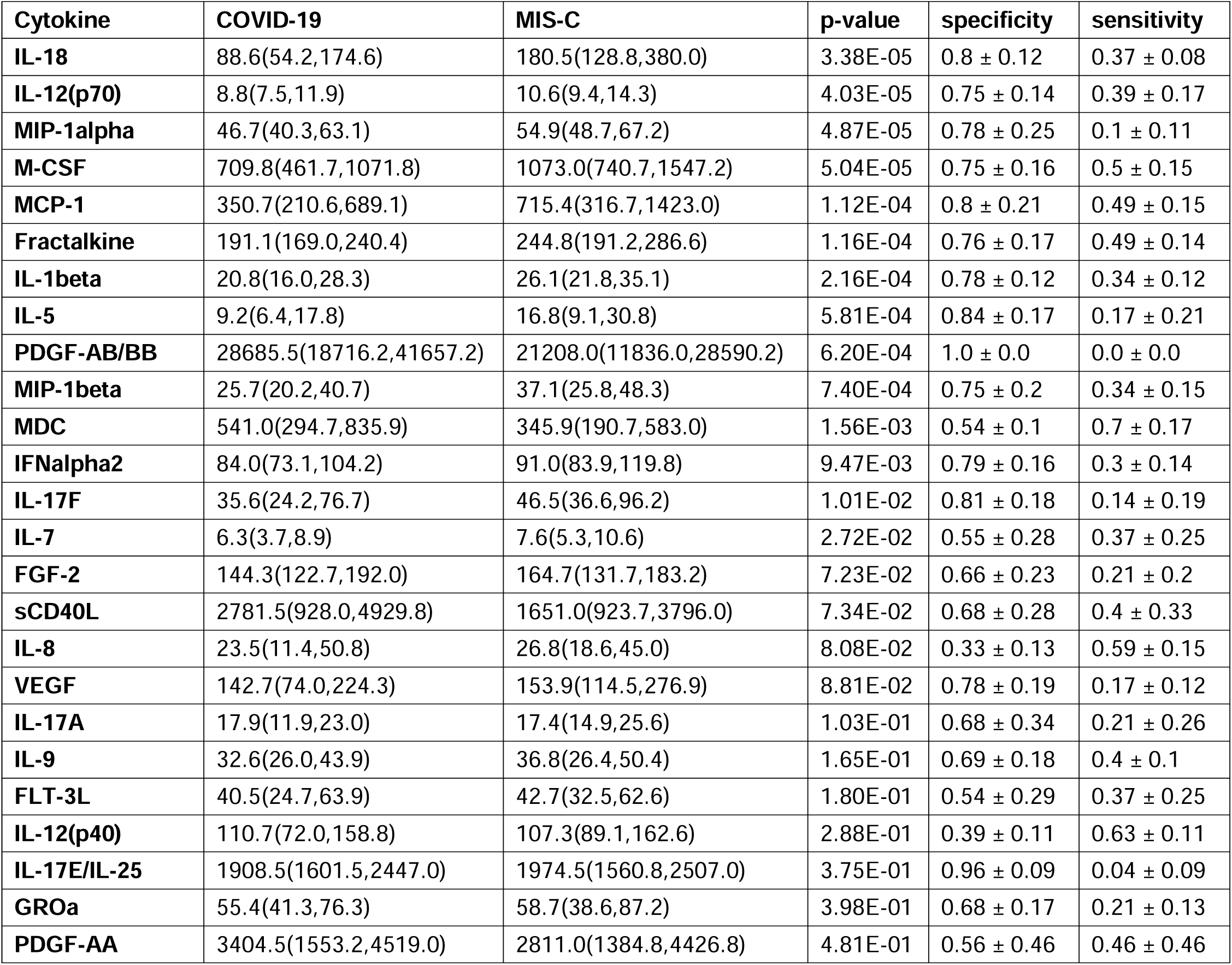
**c:** Cytokines/chemokines ordered by p-value of the Wilcoxon-rank-sum univariate test for discriminating COVID-19 from MIS- C in the training cohort (72 COVID-19, 66 MIS-C). 34 of the 45 cytokines/chemokines are statistically significant in a univariate sense. The specificity and sensitivity of each marker in a five-fold cross validation is also shown. All values are in pg/mL

### Machine learning models differentiating COVID-19 from MIS-C

Supplementary Figure 3 shows an L1 regularized logistic regression model trained by 5-fold cross-validation using the 13 standard lab biomarkers. The model selects markers, omitting Troponin I. Performance statistics of this model on the training set as well as the three validation sets are in Supplementary Table 1. On the training data, the model exhibits a 5-fold cross-validated AUC of 0.86 ± 0.05 and an F1 score of 0.78 ± 0.07. On validation set 1 of 29 COVID-19 and 43 MIS-C patients, the model makes 15 errors (5 COVID-19 and 10 MIS-C) with an AUC of 0.85 and an F1 of 0.81. On validation set 2 of 32 COVID-19 and 30 MIS-C patients, the model makes 14 errors (3 COVID-19 and 11 MIS-C) with an AUC of 0.84 and an F1 of 0.75. On validation set 3 of 20 COVID-19 and 46 MIS-C patients, the model makes 16 errors (4 COVID-19 and 12 MIS-C) with an AUC of 0.83 and an F1 of 0.71.

Figure 1 shows an L1 regularized logistic regression model trained by 5-fold cross-validation on the cytokine/chemokine data obtained from the training cohort. The model uses a total of 16 of the available 45 cytokines/chemokines to achieve a cross-validated AUC of 0.95±0.02 and F1 score of 0.91±0.04. The performance of L1-regularized models built with both cytokines/chemokines and laboratory biomarkers as predictors was identical to models built with cytokines/chemokines alone. The addition of laboratory biomarkers to the overall set of predictors was therefore determined to not improve performance of the models.

**Figure 1:**
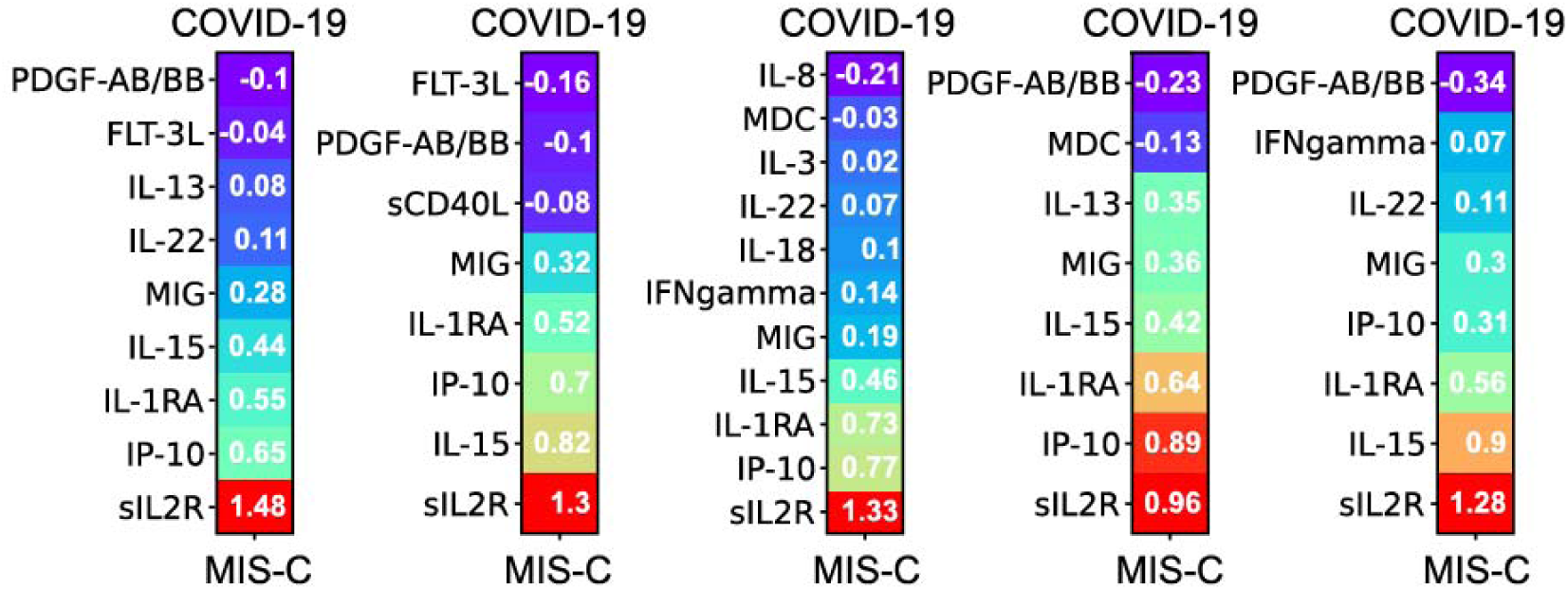
L1 regularized logistic regression model trained by 5-fold cross-validation on the cytokine/chemokine data obtained from the training cohort. The model uses a total of 16 of the available 45 cytokines/chemokines.

On validation set 1 of 29 COVID-19 and 43 MIS-C patients, the model makes 6 errors (0 COVID-19 and 6 MIS-C) with an AUC of 0.98 and an F1 of 0.93. On validation set 2 of 32 COVID-19 and 30 MIS-C patients, the model makes 8 errors (5 COVID-19 and 3 MIS-C) with an AUC of 0.89 and an F1 of 0.88. The drop in performance on the second validation set is consistent with the prevalence of the delta variant in this cohort with inflammation in COVID-19 being more severe compared to the pre-alpha training cohort. On the third validation set of 20 COVID-19 and 46 MIS-C patients, the model makes 3 errors (1 COVID-19 and 2 MIS-C) with an AUC of 0.99 and an F1 of 0.97. Importantly, these results do not change significantly even when the model is restricted to 10 cytokines/chemokines composed of the top five predictive of COVID-19 and the top five predictive of MIS-C from the original 16 biomarker model. These 10 cytokines/chemokines are soluble IL2R, IP-10 (CXCL-10), IL-1RA, IL-15, MIG (CXCL-9), MDC (CCL 22), IL-8, G-CSF, FLT-3L, and PDGF-AB/BB.

Figure 2A (top) shows a two-dimensional UMAP projection of the 45-dimensional cytokine/chemokine vector representing each sample in our training cohort. Two clusters become apparent: COVID-19 patients (purple dots) to the left and the MIS-C patients (red dots) to the right. Such separation is consistent with the excellent cross-validated performance of the logistic regression model on the training data – the two cohorts can in fact, be well separated by a linear boundary in the non-linearly projected two-dimensional UMAP coordinate frame. Note that the separation is not perfect and that the groups are not completely pure. However, the logistic regression model misclassifies only seven of them. They are MIS-C patients with a less severe form of the disease, as reflected in the cytokine/chemokine profiles shown in Supplementary Figure 4a and 4b.

**Figure 2.**
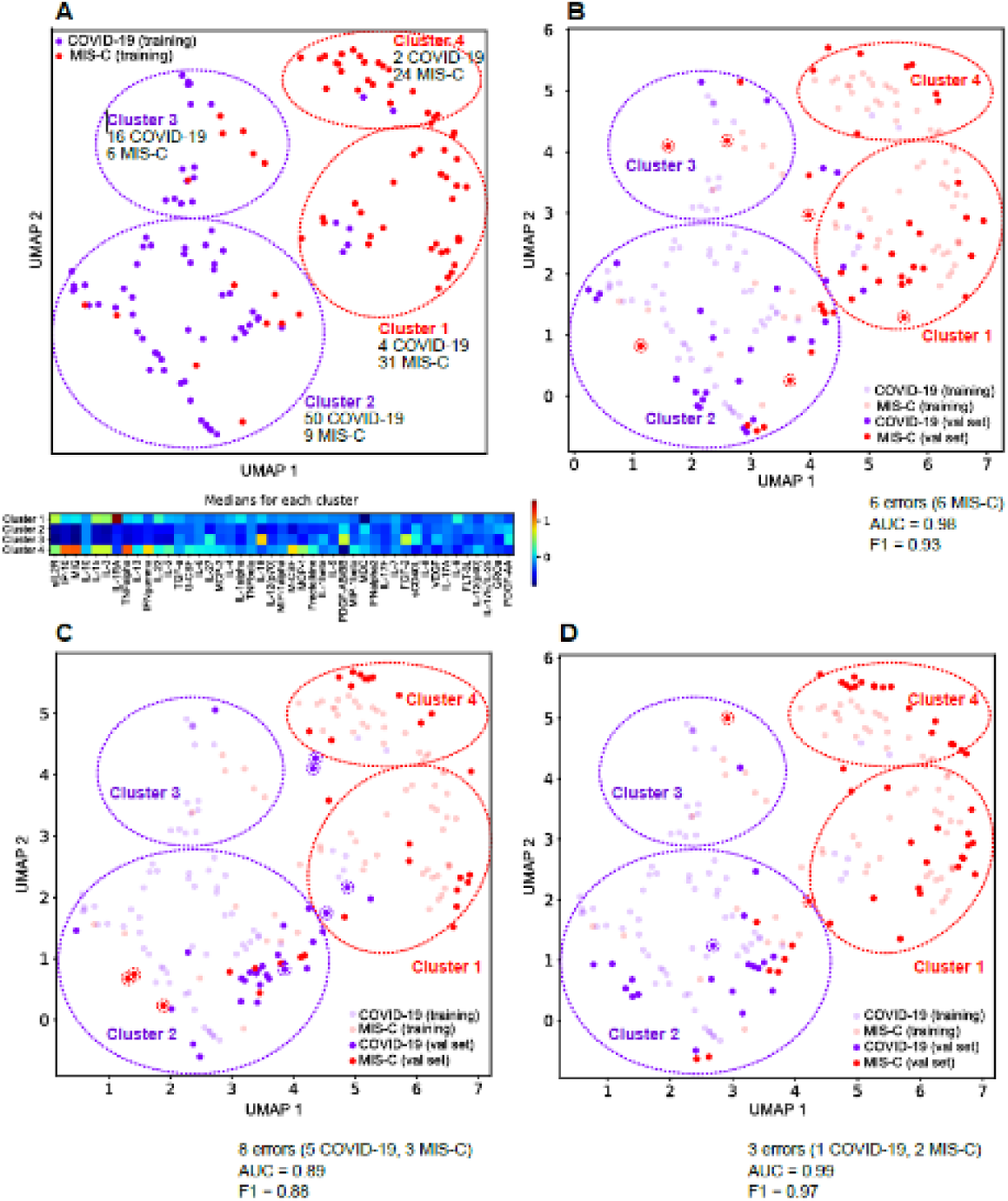
**A: Top**: a two-dimensional UMAP projection of the 45-dimensional cytokine/chemokine vector representing each asmple in our training cohort. The COVID patients in the validation set are colored purple, while the MIS-C patients are colored red.**A: Bottom**: the medians of key cytokines/chemokines for each of the four clusters.**B**: The first validation set projected back into the UMAP coordinates derived from the training data. Misclassified COVID-19 and MIS-C patients are called out by dotted circles of purple and red. For the first validation set with 29 COVID and 43 MIS-C, only 6 MIS-C patients were misclassified. 5 of them fall in the COVID clusters defined by the training cohort, and these MIS-C patients were confirmed to be mild cases by chart review.**C**: The second validation set projected back into the UMAP coordinates derived from the training data. Misclassified COVID-19 and MIS-Cpatients are called out by dotted circles of purple and red. For the second validation set with 32 COVID-19 and 30 MIS-C patients, 5COVID-19 and 3 MIS-C patients were misclassified. 4 of the COVID-19s fall in the MIS-C clusters defined by the training cohort, showing the evolution of the disease with the COVID-19 patients having more severe disease compared to the initial training cohort. The 3 misclassified MIS-C’s have a mild version of the disease and fall into the low risk COVID-19 cluster defined by the training data. **D**: The third validation set projected back into the UMAP coordinates derived from the training data. Misclassified COVID-19 and MIS-C patients are called out by dotted circles of purple and red. For the third validation set with 20 COVID and 46 MIS-C patients,1 COVID-19 and 2 MIS-C patients were misclassified.

The COVID-19 and MIS-C patients are further stratified into two clusters each. Cluster 2 (50 COVID-19, 9 MIS-C) and Cluster 3 (16 COVID-19, 6 MIS-C) are predominantly COVID-19 clusters, while Cluster 1 (4 COVID-19, 31 MIS-C) and Cluster 4 (2 COVID-19, 24 MIS-C) are predominantly MIS-C clusters. As shown in Figure 2A (bottom), Cluster 3 represents patients in the COVID-19 cohort with higher median levels of inflammation in the measured cytokines/chemokines. Relative to Cluster 2, Cluster 3 patients have elevated median levels of IL-18, IL-27, PDGF-AB/BB, and FGF-2. Cluster 4 represents patients in the MIS-C cohort with higher median levels of IP-10, MIG, TNF-a, and IFN-g, relative to Cluster 1. Cluster 1 is characterized by an elevation of IL-1RA relative to Cluster 4, potentially reflecting treatment with IVIg/steroids/anakinra. The overall risk stratification/disease severity suggested by the UMAP plot is Cluster 4 > Cluster 1 > Cluster 3 > Cluster 2 (highest to lowest risk and disease severity).

Medians and interquartile ranges for the thirteen laboratory biomarkers for each of the clusters are shown in Supplementary Table 2. As expected, the values of the lab markers in these clusters correlate well with the median cytokine/chemokine profiles from Figure 2A. In addition, we computed medians of the length of stay (days), ICU length of stay (days). Cluster 4 and Cluster 1 (the MIS-C clusters) are associated with longer stays in both the ICU and the hospital compared to Cluster 2 and Cluster 3 (the COVID-19 clusters). To assess severity of disease, we computed the fraction of patients on respiratory support (ECMO, Ventilator, CPAP) for the four clusters. Cluster 4 has the highest usage of ECMO, Ventilator and CPAP, while Cluster 2 has the least usage confirming the severity/risk ordering of the COVID-19 and MIS-C patients.

### Generalizability of model to new validation sets

Table 2 shows that the model trained on the initial cohort of 72 COVID-19 and 66 MIS-C patients performs well on 3 new validation sets from local patients and a fourth set from other sites. To understand the classification errors made by the model, we project each of the validation sets into the same UMAP coordinate frame.

**Table 2:** Performance of the logistic regression model trained on cytokines/chemokines of the training cohort and tested on three denovo validation sets gathered as the virus evolved in time, and the External Dataset. Note that the training set performance is judged by five-fold cross-validation, and thus there is a mean and standard deviation associated with each performance measure. The validation sets are evaluated in a standard train/test configuration. A single model is built with all the training data, and the validation sets are evaluated in turn against this model. Hence there is a single number characterizing the performance of the model along each metric.

|  | **AUC** | **F1** | **AUPRC** | **Accuracy** |
| --- | --- | --- | --- | --- |
| **Training set (72 C, 66 M)** | 0.95 ± 0.02 | 0.91 ± 0.04 | 0.97 ± 0.01 | 0.92 ± 0.04 |
| **Val set 1 (29 C, 43 M)** | 0.98 | 0.93 | 0.99 | 6 errors (0 C, 6 M) |
| **Val set 2 (30 C, 32 M)** | 0.89 | 0.88 | 0.91 | 8 errors (5 C, 3 M) |
| **Val set 3 (20 C, 46 M)** | 0.99 | 0.97 | 0.99 | 3 errors (1 C, 2M) |

Figure 2b shows Validation set 1 projected on the training data UMAP. Of the 29 COVID and 43 MIS-C patients in this validation set, only 6 MIS-C patients were misclassified. Note that all the new COVID-19 patients fall within the COVID-19 clusters 2 and 3 defined by the training data. 5 of the 6 misclassified MIS-C’s fall in the COVID-19 clusters defined by the training cohort, and these MIS-C patients were confirmed to be mild cases by chart review.

Figure 2c shows Validation set 2 projected on the training data UMAP. Of the 32 COVID-19 and 30 MIS-C patients, 5 COVID-19 and 3 MIS-C patients were misclassified. 4 of the COVID-19s fall in the MIS-C clusters defined by the training cohort, consistent with this set containing more delta variant infected patients with severe disease compared to the initial training cohort. The 3 misclassified MIS-C’s have a mild version of the disease and fall into the low risk COVID-19 cluster defined by the training data.

Figure 2d shows Validation set 3 projected on the training data UMAP. Of the 20 COVID-19 and 46 MIS-C patients, 1 COVID-19 and 2 MIS-C patients were misclassified. An overall milder disease profile is consistent with the accuracy of this classification.

These projections of the validation sets onto the UMAP of the training data are consistent with the cytokine/chemokine measurements yielding highly accurate predictions of MIS-C even as the disease evolved. The 16 cytokine/chemokine model as well as the minimal set of 10 cytokine/chemokines appears to be robust and maintains its accuracy over time.

### Network analysis of the cytokine/chemokine training data: The cytokine profiles in MIS-C and COVID-19 are significantly different

Figure 3A shows the elevated cytokines/chemokines in COVID-19 and in MIS-C in a network where the edges denote protein-protein interactions derived from the STRING database. An edge represents either a direct or an indirect (via a longer pathway) protein-protein interaction. Both networks reflect inflammation and immune activation, but the MIS-C network is far more extensive involving many more cytokines/chemokines displaying orders of magnitude higher levels of inflammation. Compared to controls with no known inflammatory condition, the median levels of eight chemokines/cytokines in the COVID-19 patients in our training cohort exhibit a roughly two-fold elevation over healthy controls: IL-6 (2.3), GROa (2.2), MIG (2.3), IP-10 (2.0), IL-15 (2.1), IL-12 p(70) (2.6), sCD40L (2.4), and IL-22 (2.0). In contrast, compared to healthy controls, the median levels of fifteen chemokines/cytokines in the MIS-C patients are significantly elevated: IL-6 (5.2), MIG (25.4), IP-10 (56.5), IL-15 (2.8), and IL-22 (2.2), IL-1RA (106.3), IL-10 (7.7), sI L2R (7.2), IL-18 (4.0), IFN-g (2.7), VEGF (2.6), IL-27 (2.5), IL-17F (2.2), IL-8 (2.0), and TNF-α (2.0). The first five of these cytokines/chemokines are elevated in both the MIS-C and COVID-19 groups, with levels higher in MIS-C than in COVID- 19: IL-6 (2.6), MIG (12.7), IP-10 (28.7), IL-15 (1.33), and IL-22 (1.1), where the numbers in parenthesis denote the fold-change in median levels in MIS-C over that in COVID-19 patients. The other ten cytokines/chemokines are elevated only in MIS-C patients.

**Figure 3.**
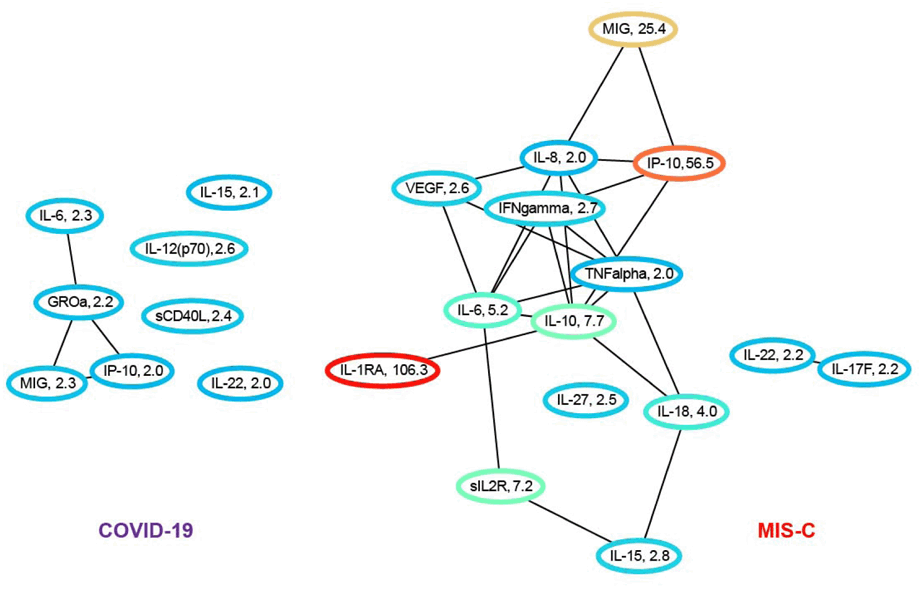
**A.** Ratio of median cytokine levels in COVID-19 patients to healthy controls and in MIS-C patients to controls for all cytokines/chemokines that are elevated/depressed two-fold or more. The network uses protein-protein interactions (black) from the STRING database to determine connections. Nodes are colored based on the fold change, and the actual fold change is also included in the node label. Many more cytokines/chemokines are inflamed in MIS-C compared to COVID-19.

In both networks, of the cytokines/chemokines we measured, IL-6, MIG, and IP-10 contribute the most to the cytokine storm. However, high levels of soluble IL2R, IL-1RA, IL-10, IL-18, IL-8, IFN-g, IL-27, IL-17F and TNF-α appear unique to MIS-C, and possibly serve as markers for disease severity. It is interesting to note that the top ten cytokines/chemokines selected by the robust L1-regularized logistic model for differentiating COVID-19 from MIS-C include MIG, IP-10, IL-15 which are three of the five cytokines/chemokines elevated in both diseases, with significantly greater elevation in MIS-C. The biomarkers sIL2R, IL-1RA, and IL-8 are elevated only in MIS-C. Of the other four biomarkers included in the model MDC (1.6) and PDGF-AB/BB (1.4) are elevated in COVID-19 relative to MIS-C, while G-CSF (2.36) and FLT-3L (1.1) are elevated in MIS-C.

To understand the role of the elevated cytokines/chemokines in the context of other unmeasured proteins, we augmented the network with proteins that interact with at least four of the measured biomarkers. The augmented networks for both COVID-19 and MIS-C are shown in Figure 3B. These longer-range interactions connect the disconnected components in both networks in Figure 4 to the core subnetworks. In the COVID-19 network in Figure 3B, IL-15, scD40L, IL-12p70 connect to the MIG/IP-10/IL-6 subnetwork via IL-4. In addition. IL-22 connects to the core subnetwork in COVID-19 in Figure 3 via IL-6. In the MIS-C network, IL-22 connects to the core connected component in Figure 3 via IL-10. Figure 3B demonstrates the essential connectedness of the networks shown in Figure 3, the extent and scope of immune system dysfunction in MIS-C, and the signaling pathways affected by the disease.

**Figure 3.**
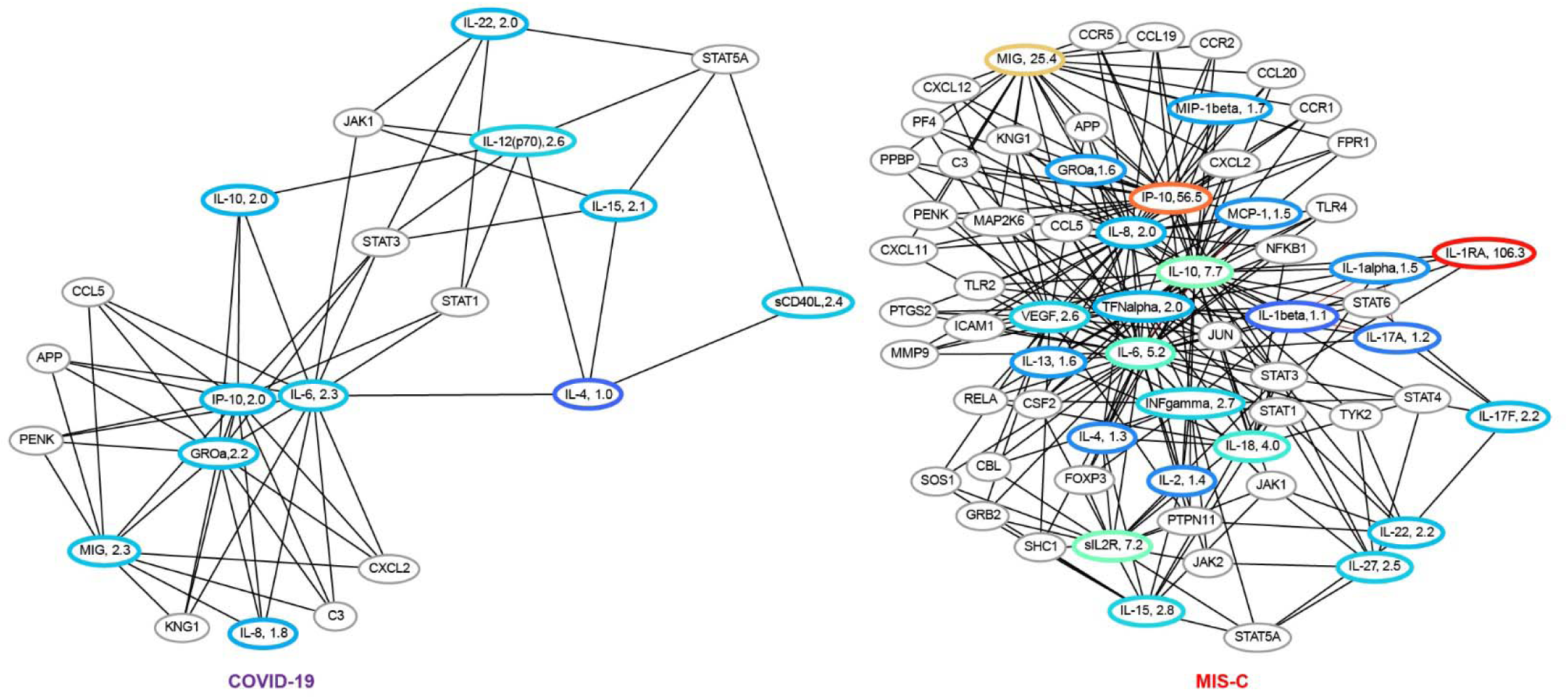
**B:** Ratio of median cytokine levels in COVID-19 and MIS-C patients to controls for all cytokines elevated or depressed at least two-fold. The network includes non-measured proteins that have protein-protein interactions with at least four measured cytokines. We can see that network dysfunction is more pronounced in MIS-C compared to COVID-19. In the COVID-19 network, we see IL-15, scD40L, IL-12 p(70) connecting to the MIG, IP-10, IL-6 subnetwork via IL-4. In addition, IL-22 connects back to core subnetwork in COVID-19 via IL-6. In the MIS-C network, IL-22 connects to the core connected component via IL-10.

Examining differential expression of the cytokines/chemokines between COVID-19 and MIS-C patients in the training cohort, Figure 3C shows cytokines/chemokines for which the ratio of the median levels in the MIS-C group to the median level in the COVID-19 group is greater than 2, i.e., a two-fold or more elevation in MIS-C. 14 cytokines/chemokines are observed to be differentially overexpressed: IL-1RA (92.7), IP-10 (27.7), MIG (11.1), sIL2R (4.2), IL-10 (3.9), IL-27 (3.2), VEGF (2.4), TNF-α (2.4), IL-18 (2.3), MCP-1 (2.3), IL-6 (2.2), IL-3 (2.1), TGF-ß(2.1), and IL-17F (2.1). The protein-protein interaction network corresponding to these differentially expressed cytokines/chemokines identifies sIL2R->IL6->IP-10->IL-10->MIG as the network path with the highest inflammation in MIS-C relative to COVID-19. These cytokines/chemokines are therefore potential targets for therapeutic intervention. Of note one of the direct branches of this pathway with extremely high differential expression (IL-1RA) in MIS-C is the target of the drug anakinra, currently used to treat this condition. Figure 3C also shows the differentially expressed cytokines/chemokines in the context of other proteins in the STRING database, revealing additional signaling pathways that could be targeted for therapeutics.

**Figure 3.**
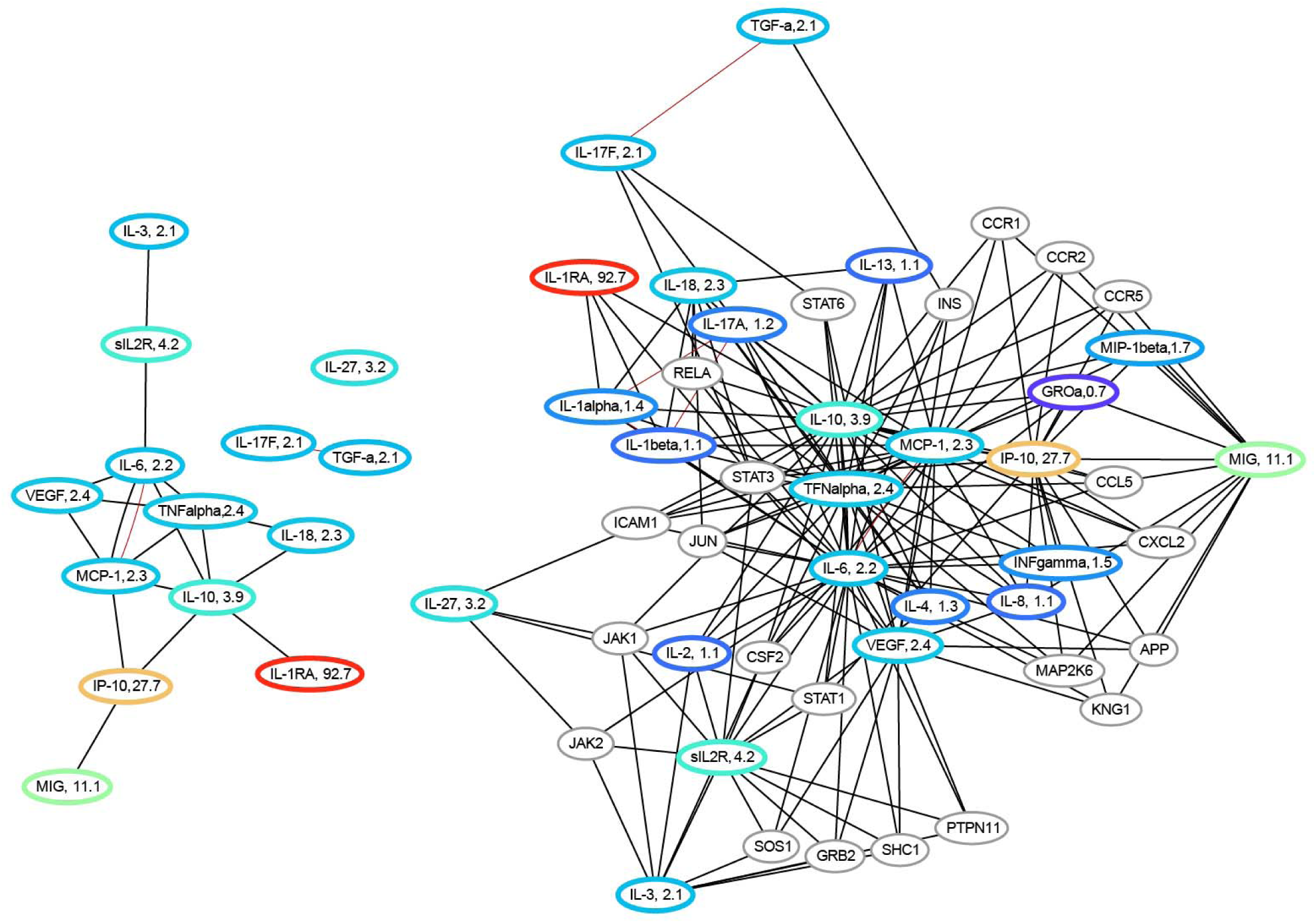
**C**: (left) Differentially expressed cytokines in MIS-C versus COVID-19 elevated or depressed at least two-fold. Nodes are labeled with the ratio of median cytokine levels in MIS-C to the median cytokine levels in COVID-19. (right) differentially expressed cytokines in context of other non-measured proteins that have interactions with at least four measured cytokines. The red edge in the left network denotes correlation of 0.8 or more in the training data between the pair of cytokines/chemokines.

## Discussion

The SARS-CoV-2 virus infects children and is typically mild in most of them, yet a small cohort develops the far more serious condition MIS-C, three or more weeks after virus exposure. This work develops and validates an objective diagnostic/prognostic method for MIS-C at initial presentation. Previous attempts to do so include a recently published meta-analysis of 787 MIS-C patients from 21 different studies (14), which identified laboratory markers including lower platelet counts, higher CRP, D-dimer, leukocyte counts, ferritin correlated with MIS-C. Our data demonstrate that standard laboratory markers of inflammation change with the severity of the variant/disease course and cannot distinguish between MIS-C and COVID-19 reliably. We therefore studied novel cytokine/chemokine markers that enable longitudinal monitoring and provide accurate patient stratification. The cytokine/chemokine markers interpreted by a comprehensive machine-learning model provide highly accurate identification of COVID-19 patients with, or at risk of developing, MIS-C. Such identification enables early intervention with IVIg/steroids. The model was trained exclusively on the May 2020 to January 2021 cytokine/chemokine data of 203 patients with COVID-19 and MIS-C, one of the largest such cohorts studied, and predicts MIS-C with a 5-fold cross-validated AUC of 0.95±0.02 and an F1 score of 0.91±0.04.

The cytokine-based model uses the levels of sixteen cytokine/chemokine markers to achieve this performance. We validated this model on three new data sets – the first with 72 patients (29 COVID-19, 43 MIS-C) seen between January 2021 and May 2021 infected predominantly of the alpha strain, the second with 62 patients (30 COVID-19, 32 MIS-C) seen between August 2021 and October 2021 with a mix of the alpha and delta strains, and the third with 66 patients (20 COVID-19 and 46 MIS-C) seen between October 2021 and January 2022, predominantly with the delta and omicron strains. The model exhibits an AUC of 0.98 and F1-score of 0.93 on the first validation data set, an AUC of 0.89 and an F1 score of 0.89 on the second validation data set, and an AUC of 0.99 and F1 score of 0.97 on the third validation dataset.

### A top ten subset of these sixteen cytokines/chemokines achieves equivalent performance on the validation data sets

AUC of 0.99 and F1 score of 0.94 on the first set, an AUC of 0.91 and an F1 score of 0.85 on the second set, and an AUC of 0.99 and an F1 score of 0.97 on the third set. These top ten cytokines/chemokines are sIL2R, IP-10, IL-1RA, IL-15, MIG, MDC, IL-8, G-CSF, FLT-3L, and PDGF-AB/BB. Our results form the basis of a new rapid multiplex assay for risk stratification of patients infected with SARS-CoV-2 virus. The ability of the model trained on the pre-alpha variant to predict MIS-C with high AUC and F1 scores on new validation sets gathered as the virus evolved affirms its robustness and generalizability to new variants as well as to new populations.

UMAP visualizations of the high-dimensional cytokine/chemokine data separates COVID-19 from MIS-C patients well, consistent with the accuracy of the L1-regularized logistic regression model. Clustering of the UMAP plots reveal finer-grained subsets, sorting MIS-C and COVID-19 patients into two groups each based on severity, providing insights into the cytokine profiles of mild vs. severe disease. By projecting the new validation data sets into the UMAP coordinate frame defined by the training data (pre-alpha strain), it is possible to track the evolution of COVID-19 and MIS-C through the progression of variants. We observe the drift of more recent MIS-C patients toward the COVID-19 training cohort clusters, and the drift of the COVID-19 patients toward the MIS-C training cohort clusters in Figures 2b, 2c and 2d, reflecting the changes in median laboratory biomarker values in the three validation sets, reported in Table 1b. The clusters derived from the UMAP visualizations significantly correlated to the severity of disease, with Cluster 4 having the highest usage of ECMO, Ventilator and CPAP, while Cluster 2 has the least usage confirming the severity/risk ordering of the COVID-19 and MIS-C patients.

The sixteen-cytokine/chemokine panel as well as the top ten subset generate useful system-level hypotheses about the pathogenesis of MIS-C. By deriving a protein-protein network using the STRING database, with elevated cytokines/chemokines in COVID-19 and MIS-C as nodes, we visualize the affected signaling pathways in both these conditions. Network analysis of the cytokines and chemokines elevated in COVID-19 versus MIS-C reveals major differences in the scope of the inflammatory response. Eight of the 45 measured cytokines/chemokines are elevated in COVID-19 and fifteen are elevated in MIS-C. Both networks show immune system dysfunction, but the MIS-C network is far more extensive, involving many more cytokines/chemokines displaying orders of magnitude higher levels of inflammation. In both networks, IL-6, MIG, and IP-10 contribute the most to the cytokine storm. However, high levels of soluble IL2R, IL-1RA, IL-10, IL-18, IL-8, IFN-1, IL-27, IL-17F and TNF-α appear unique to MIS-C and could serve as markers for disease severity.

The immunological features of pediatric COVID-19 and MIS-C are the subject of numerous investigations (15-25). Our patient cohort is one of the largest that been studied to date, and our set of 45 cytokine/chemokines is one of the most comprehensive panels to be analyzed. Amongst the different investigators who have studied differences between COVID and related diseases and MIS-C, one of the first to report clear differences between KD and MIS-C using IL-6, IP-10 and IL-17A was a small study (25) of 13 MIS-C patients. Recently other studies (16-19, 26-28), have also shown differential cytokine/chemokine expression in MIS-C compared to either controls, COVID-19 or Kawasaki disease. In a small cohort (29) of 7 MIS-C patients, differentiation between COVID-19 and MIS-C patients was achieved using a combination of IL- 10, IL-1RA, IL-18, IL-6, TNF and IFN-gamma. Subsequently (30) in a larger cohort of 118 subjects, significant elevations in IL-6, IL-10, IL-17A and IFN-gamma were demonstrated, and correlated to length of hospital stay.

Network analysis reveals that the top ten cytokines/chemokines selected by the robust L1-regularized logistic model for differentiating COVID-19 from MIS-C include a subset (MIG, IP-10, IL-15) which are three of the five cytokines/chemokines elevated in both conditions, with significantly greater elevation in MIS-C. The biomarkers sIL2R, IL-1RA, and IL-8 are elevated only in MIS-C. Of the other four biomarkers included in the model MDC (1.6) and PDGF-AB/BB (1.4) are elevated in COVID-19 relative to MIS-C, while G-CSF (2.36) and FLT-3L (1.1) are elevated in MIS-C, consistent with the model’s robust performance across a range of de novo validation sets gathered even as the disease itself evolved. The L1-regularized logistic regression model based on the measurement of as few as 10 novel cytokine/chemokine analytes provides a highly sensitive and specific method to predict MIS-C at initial presentation of SARS-CoV-2 infected patients, while also providing key insights into potential therapeutic targets.

## Conclusions

Several investigations have examined different laboratory markers of inflammation, disease activity and cytokine/chemokine signatures for MIS-C and COVID-19. These studies suggest that there is no single laboratory or cytokine/chemokine biomarker that can differentiate MIS-C and COVID-19. This motivated our quest for a multianalyte profile with algorithmic interpretation. Our model which was derived from a pre-alpha strain training cohort is accurate in predicting disease status for all major COVID variants to date (alpha, delta, omicron) in large validation cohorts and identifies MIS-C with few errors. With the change in the course and severity of the disease as well as its management with time, it is important to note that standard laboratory markers do not add any significant information to our model. Notably, the model appears to work for both diagnosis and prognosis. Roughly half of the MIS-C patients in this study had not received a MIS-C diagnosis at the time of blood sampling because they had not (yet) met the CDC criteria for MIS-C. In these patients, the model was therefore a prognostic indicator, *predicting their diagnosis at a future date*. This is important especially because with the advent of the vaccines, many of these MIS-C patients that were diagnosed based on presence of SARS-CoV-2 Spike antibodies per definition, can no longer be differentiated by presence of these antibodies since they also mount a robust post-vaccine antibody response.

Unique in our cohorts is the time interval between each subset of patients, reflecting distinct variants of the SARS CoV-2 virus in those patients, and that our 10-biomarker model provides a highly sensitive and specific method to predict MIS- C at initial presentation of SARS-CoV-2 infected patients. We elucidate plausible pathways for immune dysregulation in MIS-C. A protein-protein interaction network analysis identifies sIL2R, IL6, IP-10, IL-10, MIG as the network path with the highest inflammation in MIS-C relative to COVID-19, indicating that there are two pathways, mediated by NFkB and IFN- 1 leading to elevated IP-10 and MIG. Understanding the interaction of these pathways as it relates to disease severity in pediatric MIS-C patients is key to providing insights into potential therapeutic targets, as differential responses to SARS-CoV-2 vaccines in this population emerge.

## Study Approval

Institutional review Board at Baylor College of Medicine

## Data Availability

All data produced in the present study are available upon reasonable request to the authors

## Acknowledgements

NIH (Grant R61HD105593). Karen Prince for artistic rendition of figures, Deepthi Rajapakshe for sample processing.

---

**Supplementary Figure 1.**
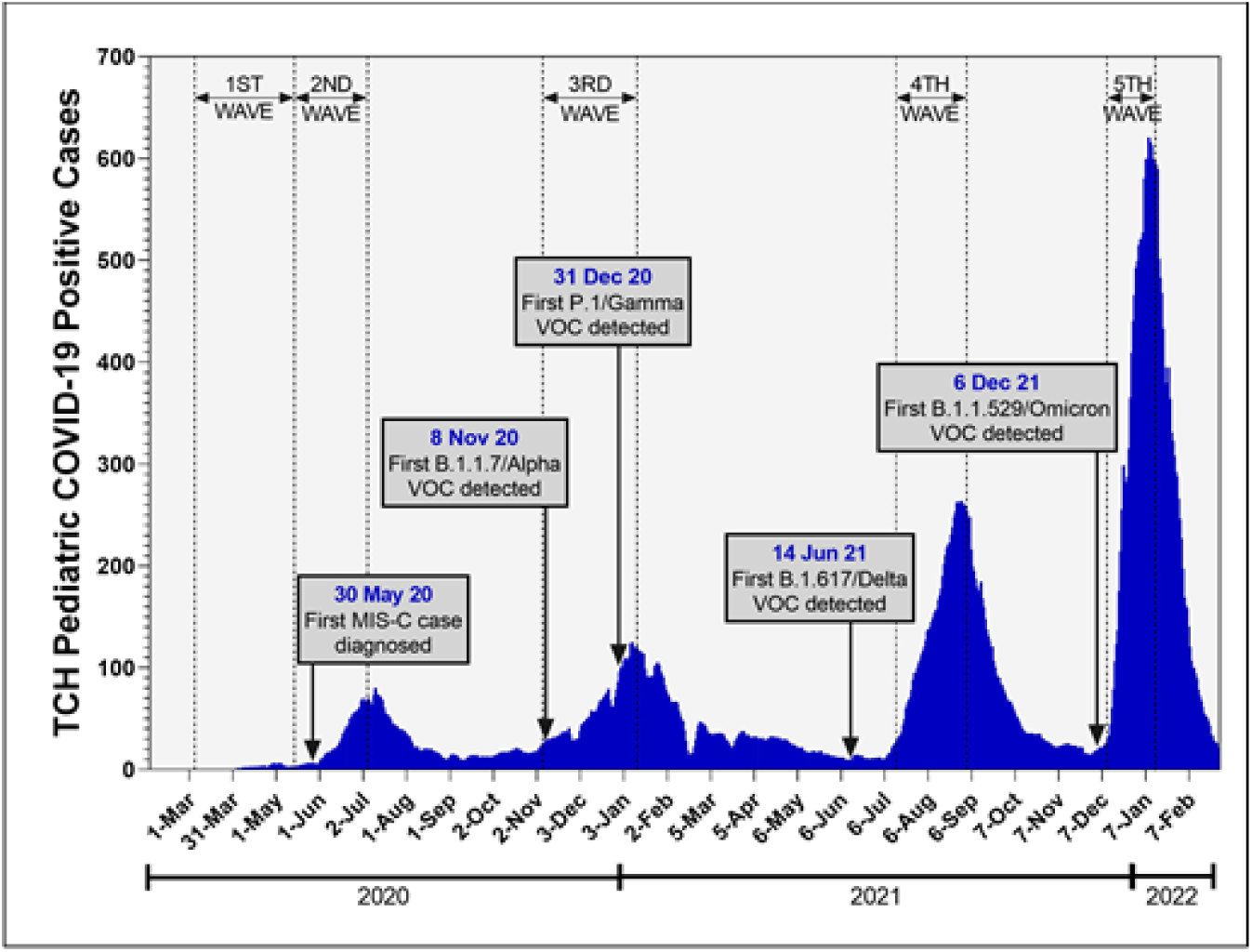
**a:** SARS-CoV-2 positive case trends at TCH during the COVID-19 pandemic. These data represent the 7-day average of SARS-CoV-2 positive cases over time from March 2020 – February 2022. Specimens from positive cases have been stored at TCH-CB for future studies.

**Supplementary Figure 1.**
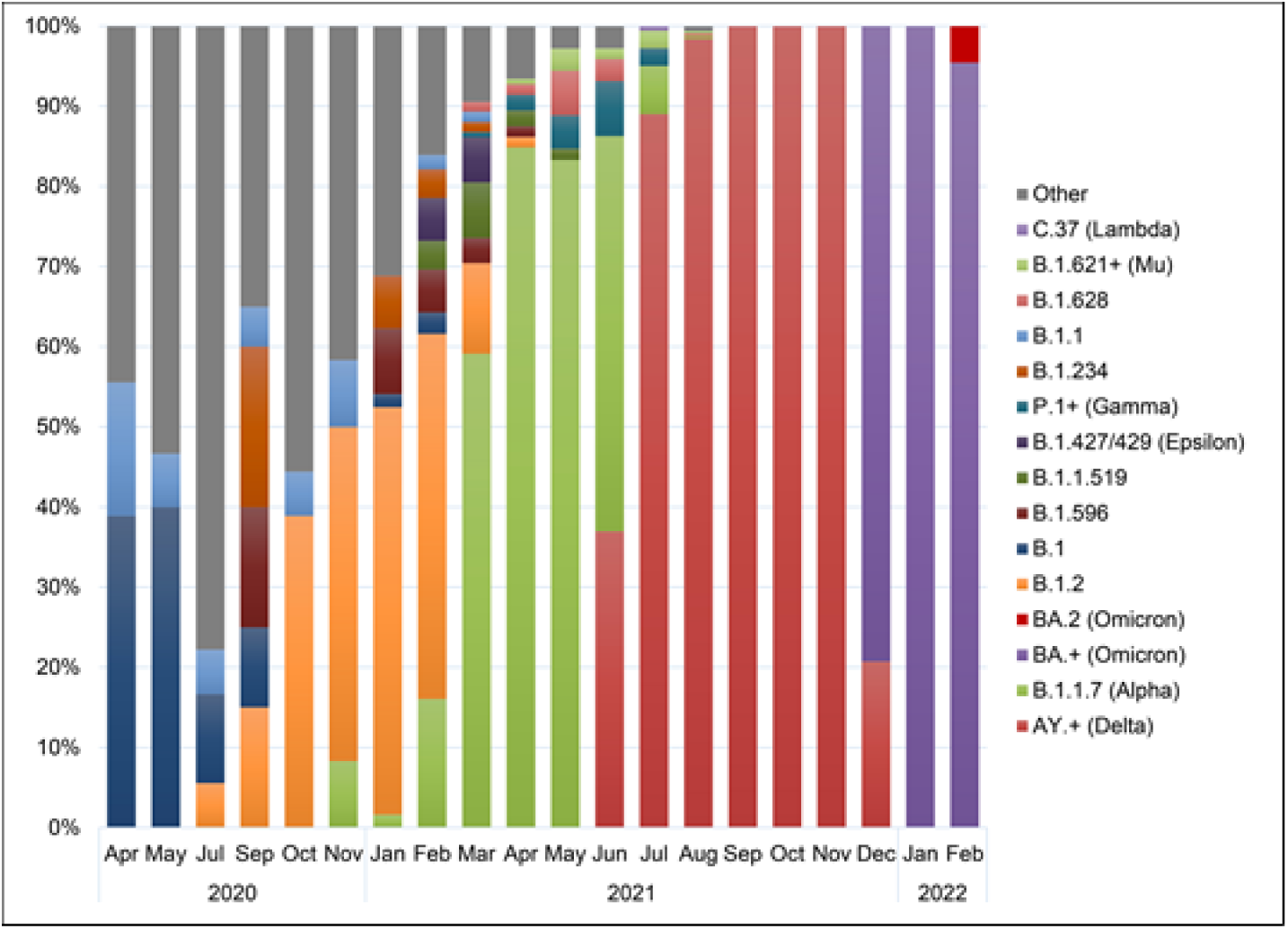
**b:** Temporal changes in circulating SARS-CoV-2 variants in pediatric patients. Significant changes have been observed in SARS-CoV-2 variants identified at TCH over the past 22 months. Predominance of lineages have shifted from B.1 in 2020 and B.1.2 in January 2021 to a rapidly shifting variant profile with the emergence of VOCs B.1.1.7 (Alpha), B.1.617/AY.+ (Delta), and BA.+ (Omicron).

**Supplementary Figure 2:**
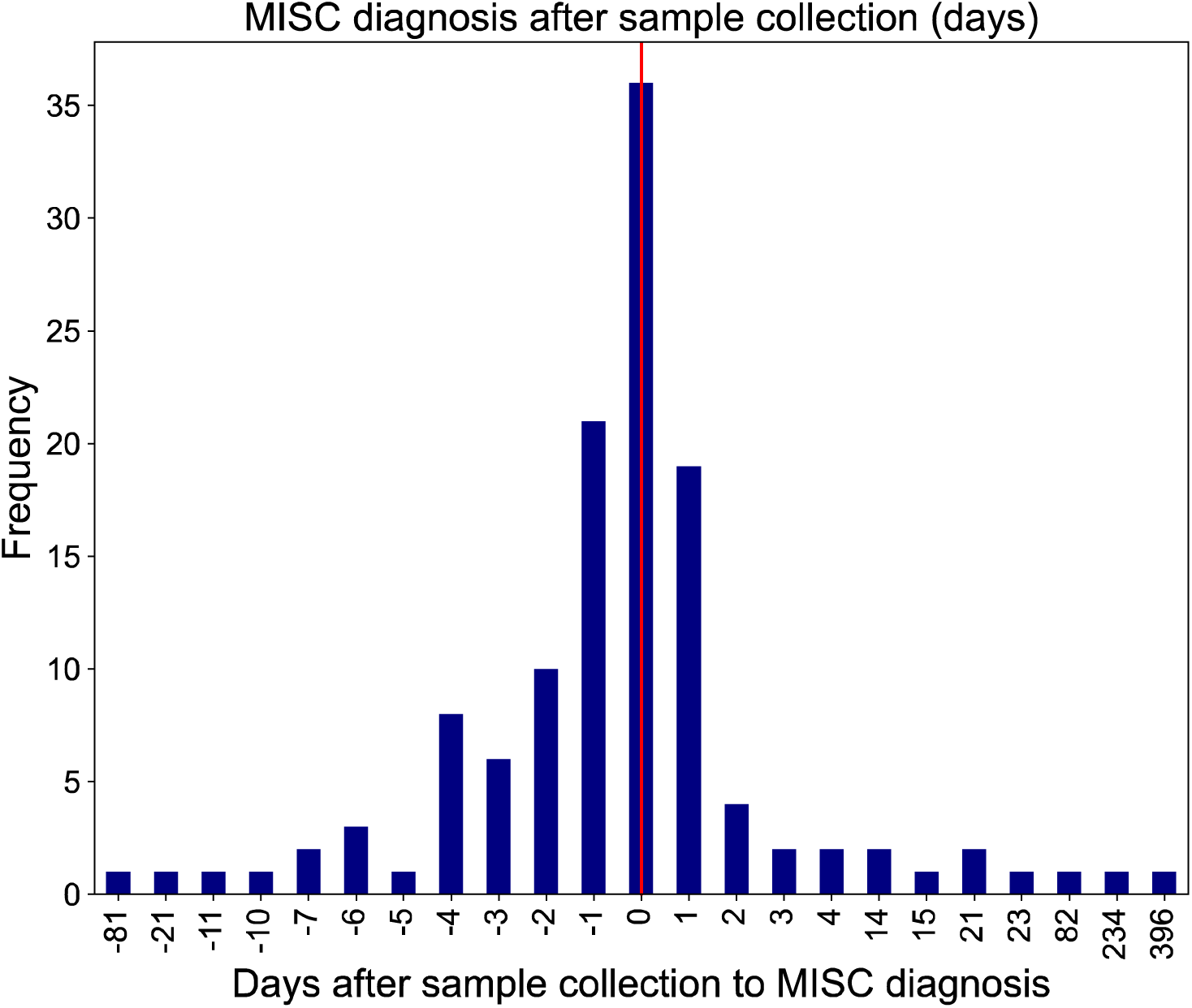
Distribution of the number of days after sample collection that a diagnosis of MIS-C was made in the MIS-C cohort in the training set, and validation sets 1 and 2. Negative numbers on the x-axis indicates the sample collection was made after the diagnosis of MIS-C was called. More than half of the MIS-C group had their samples collected prior to diagnosis.

**Supplementary Figure 3:**
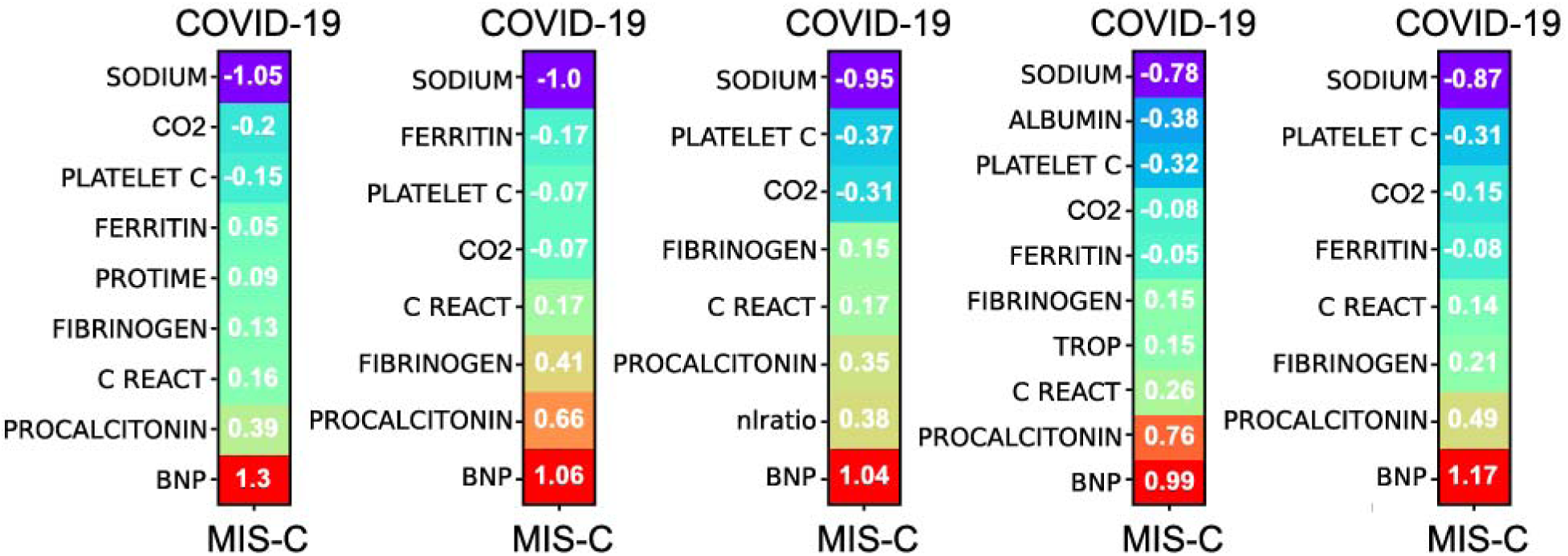
5-fold cross-validated L1 regularized model logistic trained by cross-validation using lab biomarker data only. The model uses a total of 12 lab biomarkers, note that the model does not select Troponin I.

**Supplementary Figure 4.**
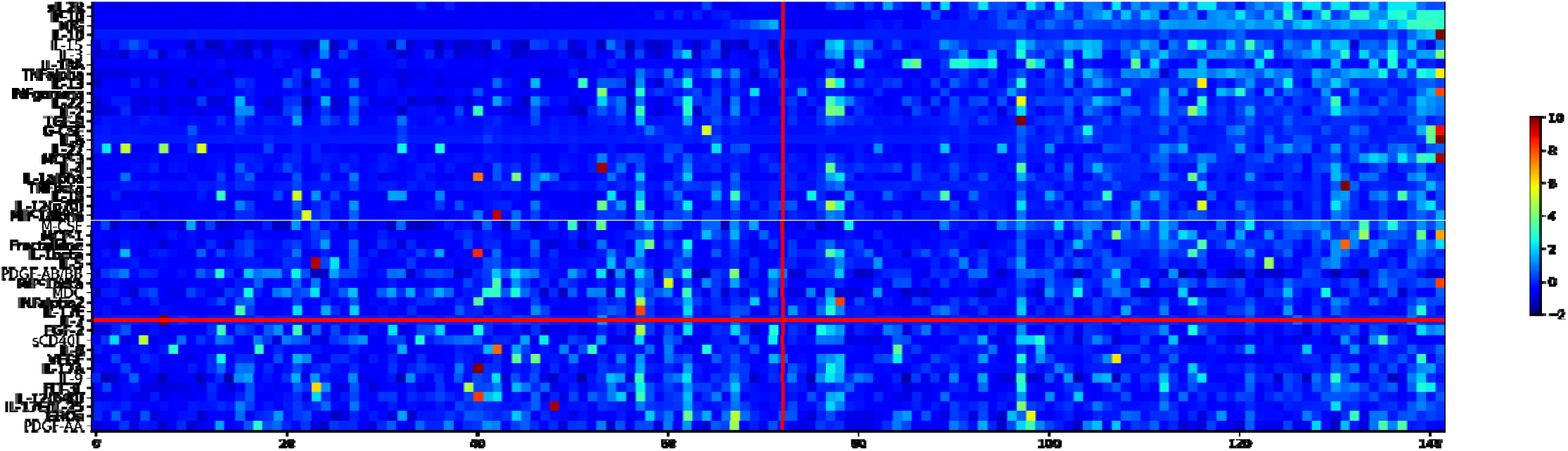
**a:** Sorted heat map of 45 measured cytokines/chemokines for the MIS-C and COVID-19 patients in the training set. Each column represents the chemokine/cytokine profile for a patient. The color gradient represents the standardized values of each of the cytokines/chemokines. Patients to the left of the red vertical line are COVID-19 patients, while those ot the right are MIS-C. The MIS-C cohort has elevated levels of cytokines/chemokines on average, although the COVID-19 cohort also has patients with elevated levels at the right end. The cytokines/chemokines on the y-axis are sorted in ascending order basedon the p- value of the Wilcoxon-rank-sum test to differentiate COVID-19 samples from MIS-C. The horizontal red line demarcates those cytokines/chemokines whose p-value falls above 0.05.

**Supplementary Figure 4.**
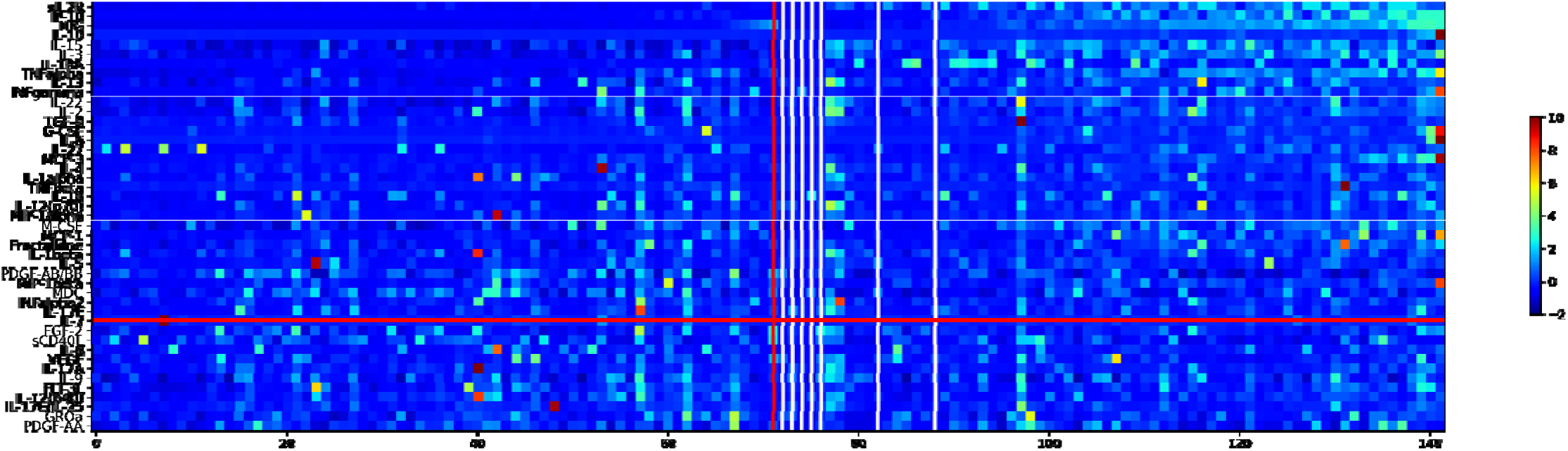
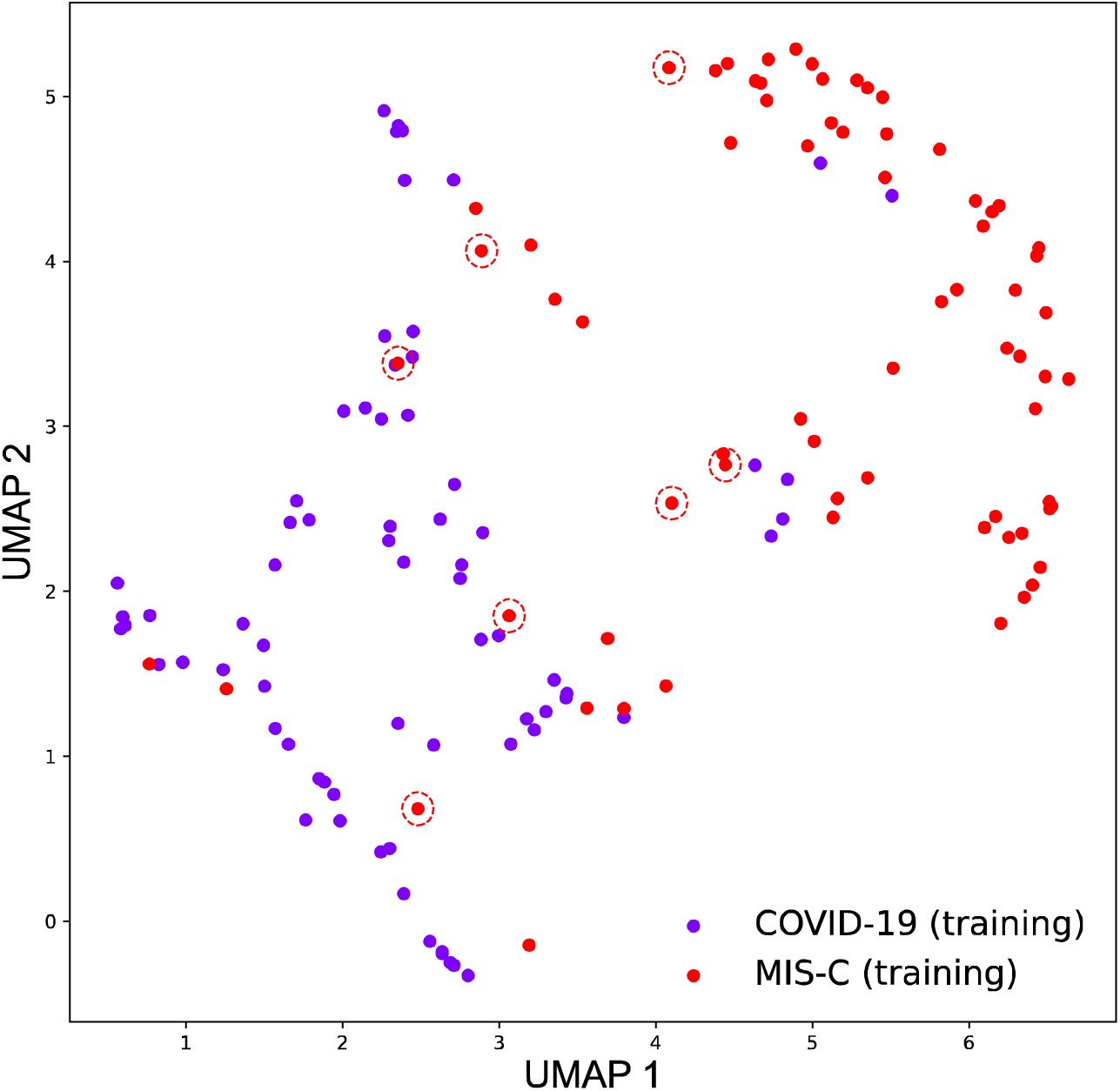
**b: Top:** Sorted heat map of 45 measured cytokines/chemokines for the MIS-C and COVID-19 patients in the training set, with classification errors made by the model on the training set itself. Each column represents the chemokinec/ytokine profile for a patient. The color gradient represents the standardized values of each of the cytokines/chemokines. Patients ot the left of the red vertical line are COVID-19 patients, while those to the right are MIS-C. Note that all seven errors are MIS-C patients with low inflammation levels. **Bottom:** The classification errors in the training set projected into the UMAP space. Note that all s ven errors are MIS-C patients, and that six of them map into the COVID-19 clusters, indicating lower than expected inflammation levels, resembling COVID-19 rather than MIS-C.

**Supplementary Table 1:** Performance of logistic regression model trained on laboratory biomarkers of the initial cohort and tested on three denovo validation sets.

|  | **AUC** | **F1** | **AUPRC** | **Accuracy** |
| --- | --- | --- | --- | --- |
| **Training set (72 C, 66 M)** | 0.86 ± 0.05 | 0.78 ± 0.07 | 0.88 ± 0.06 | 0.81 ± 0.06 |
| **Val set 1 (29 C, 43 M)** | 0.85 | 0.81 | 0.89 | 15 errors (5 C, 10 M) |
| **Val set 2 (30 C, 32 M)** | 0.84 | 0.75 | 0.84 | 14 errors (3 C, 11 M) |
| **Val set 3 (20 C, 46M)** | 0.83 | 0.71 | 0.86 | 16 errors (4 C, 12 M) |

**Supplementary Table 2:** Characterizing cytokine/chemokine derived UMAP clusters by lab and hospital data. Medians and interquartile ranges are shown for the lab markers. Cluster 2 and Cluster 3 contain predominantly COVID-19 patients, while Clutser 1 and Cluster 4 are primarily MIS-C.

|  | **Cluster 1** | **Cluster 2** | **Cluster 3** | **Cluster 4** |
| --- | --- | --- | --- | --- |
| **CRP** | 7.3 (2.6,18.1) | 3.2 (0.9,7.4) | 6.3 (0.8,16.9) | 16.4 (4.3,22.2) |
| **Procalcitonin** |  |  |  |  |
| **D-Dimer** | 2.1 (1.3,3.5) | 1.3 (0.6,2.6) | 1.7 (0.5,3.9) | 3.9 (2.1,5.8) |
| **BNP** | 109.4 (50.0,342.9) | 63.8 (17.9,68.3) | 66.8 (29.6,138.7) | 300.1 (65.0, 706.2) |
| **Sodium** | 134 (132,139) | 138 (135,140) | 136.5 (133.3,138) | 134.5 (131.3, 137) |
| **Platelet counts** | 207 (119.5,277.5) | 243 (161.5,396) | 181 (149.8,313.5) | 166 (144.3,241) |
| **Albumin** | 3.3 (2.9,3.8) | 4.0 (3.4,4.6) | 3.5 (3.2,4.2) | 3.4 (3.1,3.6) |
| **Fibrinogen** | 458 (383.8, 558) | 406.3 (325,535.5) | 449 (389.5,596) | 485 (385.9,583.8) |
| **Prottime** | 15.3 (14.2,16.5) | 14.8 (14,15.2) | 14.9 (14.7,16.1) | 15.3 (14.5,16.2) |
| **NL ratio** | 9.1 (3.5,14.7) | 2.4 (1.3,5.4) | 5.3 (2.7,11.0) | 6.1 (1.9,14.6) |
| **CO<sub>2</sub>** | 24 (21,28) | 26 (23,27) | 26 (24,27.8) | 23 (20,26) |
| **Ferritin** | 327.2 (159,485.5) | 254.0 (117.5,379.7) | 324.4 (147,507.2) | 242 (124.3,649.0) |
| **Troponin I** | 0.04 (0.01, 0.11) | 0.02 (0.01, 0.07) | 0.02 (0.01,0.07) | 0.02 (0.01,0.07) |
| **Length of stay (days) med** | 7.9 | 6.7 | 5.2 | 7.82 |
| **ICU LOS (days) median** | 3.7 | 0 | 2.4 | 4.47 |
| **ECMO (%)** | 0.0 | 2.1 | 0.0 | 10.7 |
| **CPAP (%)** | 11.1 | 17.0 | 25.0 | 32.1 |
| **Ventilator (%)** | 8.3 | 23.4 | 33.3 | 35.7 |
| **AKI (%)** | 11.1 | 12.8 | 45.8 | 35.7 |
| **IVIg/steroids 7dys (%)** | 38.9 | 34.0 | 50.0 | 39.3 |

## Notes

### Competing Interest Statement

The authors have declared no competing interest.

### Funding Statement

NIH R61-HD 105593

### Author Declarations

IRB Committee of Baylor College of Medicine gave ethical approval of this work

